# Sex-Related Differences in Eicosanoid Levels in Chronic Thromboembolic Pulmonary Hypertension

**DOI:** 10.1101/2022.05.20.22274927

**Authors:** Mona Alotaibi, Timothy Fernandes, Amber B. Tang, Gino Alberto Magalang, Jeramie D. Watrous, Tao Long, Victor Pretorius, Michael Madani, Nick H. Kim, Mohit Jain, Susan Cheng

## Abstract

A sex paradox is seen in pulmonary arterial hypertension (PAH) whereby female sex is a risk factor for development of the disease but male sex is often associated with poorer outcomes. Although data are limited regarding the potential molecular mechanisms underlying sex-specific variations in PAH including its various subtypes, there is emerging evidence regarding the potential role of sex differences in pathways governing the balance between underlying inflammatory and vasoactive mechanisms. Chronic thromboembolic pulmonary hypertension (CTEPH) is a subtype of pulmonary hypertension that is classically characterized as a procoagulant state and yet also involving some degree of proinflammatory activity. Therefore, we sought to determine whether eicosanoid mediators, endogenous lipid signaling molecules that have been implicated in a range of inflammatory and vasoactive responses, vary by sex in patients with CTEPH. We studied 287 patients (43.5% female) with confirmed CTEPH from a single center and assayed plasma samples using liquid chromatography – mass spectrometry to profile 500 eicosanoid analytes. We found significant sex-specific variations in eicosanoid profiles after adjusting for age and BMI, with males demonstrating a predominance of inflammatory-pathway related eicosanoids. Sex-specific variation in eicosanoid pathway related mechanisms may underlie sexual dimorphism in CTEPH outcomes.

## INTRODUCTION

Emerging data have revealed a persistent sex paradox in pulmonary arterial hypertension (PAH). Whereas women have greater risk for developing the disease than men, women go on to consistently experience better functional outcomes and survival even after accounting for differences in age, comorbidities, and interventions.^1,2^ This paradox has been attributed to sex hormonal differences between males and females, but sex variations in the underlying molecular pathophysiology have not been fully elucidated.^3^ Amidst the overall wide range of pulmonary hypertension (PH) phenotypes, the distinguishing characteristics of CTEPH (classified as group 4 PH) include chronic thrombi in the pulmonary arteries leading to increased pulmonary vascular resistance (PVR). Although CTEPH is classically characterized as a procoagulant state, underlying mechanisms are also known to involve some degree of proinflammatory activity.^4^ Notably, endogenous bioactive lipid mediators, known as eicosanoids, are recognized as not only key molecular drivers of the inflammatory and vasoactive response to stress and injury, but are also implicated in PH^5,6^ overall as well as PAH and CTEPH,^7^ in particular. Intriguingly, eicosanoids are also known to exhibit sexual dimorphism^8^ and thus sex-specific variation in eicosanoid pathways could represent an important mechanism contributing to sex differences in PAH including CTEPH.

## METHODS

### Study Cohort

Patients were enrolled if they have confirmed diagnosis of CTEPH and were scheduled for PEA surgery at the University of California San Diego (UCSD) between 1/2014 and 12/2018. Diagnosis of PH was confirmed by standard criteria on right heart catheterization (RHC).^9^ CTEPH was confirmed based on results of ventilation/perfusion scan and computed tomography scan or pulmonary angiography performed ≥3 months after uninterrupted anticoagulation therapy. All patients had fasting plasma samples collected prior to PEA surgery. Samples stored in the UCSD biobank. All study protocols were approved by the UCSD institutional review board and all patients signed informed consent.

### Eicosanoids Profiling

For all patients, plasma samples underwent liquid chromatography – mass spectrometry (LC-MS) based eicosanoids profiling using methods described previously.^10^ Following Qc/Qa analysis, data were extracted using image processing and machine learning based spectral optimization,^10^ then normalized, aligned, and filtered for statistical analyses.

### Statistical Analyses

Prior to all statistical analyses, eicosanoid analyte values were natural logarithmically transformed, then standardized (mean=0, SD=1) to facilitate inter-analyte comparisons of associations. Between-group comparisons were performed using t-test or the Mann-Whitney test for continuous variables and the χ^2^ test for categorical variables. We used logistic regression analysis to identify eicosanoids associated with male versus female in models adjusting for age and body mass index (BMI). We considered a Bonferroni corrected *P* value significance threshold of 0.05 divided by a conservative estimate of the total number of unique small molecules (i.e., *p*<10^−4^). All statistical analyses were performed using R v3.5.1.

## RESULTS

The study cohort included 287 patients with CTEPH, mean age of 55 years, of which 125 (43.5%) were female. Table 1 shows demographics and clinical characteristics for male and female patient populations. Overall, we observed similar between-group characteristics for the male and female cohorts, including age, pulmonary hypertension medications, functional class, and UCSD surgical level of disease. Among both males and females, 55% of patients were taking pulmonary hypertension medications, the most common of which was Riociguat. Both male and female patients were most commonly functional class 3 (64.2% of males and 69.6% of females). Of note, females had a slightly elevated BMI compared to males (31.5 versus 29.3, *p*=0.013), while males had a significantly higher prevalence of coronary artery bypass graft surgery (14.8% versus 4.0%, *p*=0.005). The hemodynamic profile before PEA was overall similar among males and females with no significant difference in mean right atrial pressure, pulmonary arterial pressures, and pulmonary vascular resistance between sexes (**Table 2**). Males had higher concomitant CABG surgery than females. Post-operative hemodynamics were similar except for men had better PVR and CO (*p*= 0.03 and <0.001 respectively).

**Table 1.** Patient cohort characteristics.

|  | **Male** | **Female** | **p-value** |
| --- | --- | --- | --- |
| n | 162 | 125 |  |
| Age, years | 56.3 (15.1) | 53.9 (14.0) | 0.17 |
| Body mass index, kg/m<sup>2</sup> | 29.3 (6.2) | 31.4 (8.1) | 0.01 |
| PH-targeted therapy, (%) | 90 (55.6) | 69 (55.2) | 1.00 |
| Soluble guanylate cyclase stimulator | 62 (54.4) | 48 (60.0) | 0.53 |
| Phosphodiesterase-5 inhibitor | 21 (21.4) | 17 (23.3) | 0.92 |
| Endothelin receptor antagonist | 11 (11.7) | 17 (23.3) | 0.08 |
| Prostacyclin | 9 (9.9) | 6 (9.1) | 1.00 |
| Inhaled prostacyclin therapies | 0 (0.0) | 5 (7.5) | 0.03 |
| Functional Class |  |  | 0.84 |
| 1 | 1 (0.6) | 2 (1.6) |  |
| 2 | 45 (27.8) | 29 (23.2) |  |
| 3 | 110 (67.9) | 90 (72) |  |
| 4 | 6 (3.7) | 4 (3.2) |  |
| Coronary artery bypass graft (%) | 24 (14.8) | 5 (4.0) | 0.01 |
| Valve repair (%) | 5 (3.1) | 4 (3.2) | 1.00 |
| Patent foramen ovale closure (%) | 33 (20.4) | 29 (23.2) | 0.67 |
| Total cardiopulmonary bypass, minutes | 260.7 (40.31) | 241.3 (31.2) | <0.001 |
| Surgical classification: Level Right, (%) |  |  | 0.06 |
| 0 | 0 (0.0) | 1 (0.8) |  |
| 1 | 40 (25.0) | 15 (12.0) |  |
| 1C | 0 (0.0) | 1 (0.8) |  |
| 2 | 74 (46.2) | 62 (49.6) |  |
| 3 | 35 (21.8) | 38 (30.4) |  |
| 4 | 11 (6.9) | 8 (6.4) |  |
| Surgical classification: Level Left, (%) |  |  | 0.32 |
| 0 | 2 (1.2) | 3 (2.4) |  |
| 1 | 19 (11.9) | 7 (5.6) |  |
| 2 | 60 (37.5) | 51 (40.8) |  |
| 3 | 61 (38.1) | 44 (35.2) |  |
| 4 | 18 (11.2) | 20 (16.0) |  |
| Mechanical ventilation, days | 4.11 (6.52) | 2.18 (3.32) | 0.01 |
| Intensive care unit length-of-stay, days | 6.38 (6.57) | 4.50 (3.06) | 0.01 |
| Postoperative length-of-stay, days | 12.54 (7.97) | 10.19 (3.47) | 0.01 |
| Total length-of-stay, days | 13.81 (8.17) | 11.34 (3.86) | 0.01 |
| Reperfusion injury, (%) | 31 (19.1) | 22 (17.7) | 0.88 |
| Post operative PVR >400 dyn·s/cm<sup>5</sup>,(%) | 16 (9.9) | 9 (7.2) | 0.6 |

*Values are reported as mean and standard deviation unless indicated otherwise.*
*Abbreviations:* PH: pulmonary hypertension. Surgical classification was based on the UCSD surgical classification system.<sup>13</sup> UCSD level I: lesions starting in the main pulmonary arteries; level II: lesions starting at the level of the lobar or intermediate pulmonary arteries; level III: lesions at the level of segmental arteries only; and level IV: lesions starting at the subsegmental branches only.

**Table 2.** Patient cohort pre- and post-pulmonary endarterectomy hemodynamic measures.

|  | Male | Female | *p*-value |
| --- | --- | --- | --- |
| ***Pre-operative hemodynamics*** |  |  |  |
| RAP, mm Hg | **10.0 (7.1)** | **8.9 (5.4)** | **0.15** |
| mPAP, mm Hg | 42.4 (10.8) | 41.1 (11.5) | 0.34 |
| CO, L/min | 4.7 (1.3) | 4.7 (1.3) | 0.88 |
| CI, L/min/m | 2.2 (0.5) | 2.4 (0.6) | <0.001 |
| PVR, dyn·s/cm<sup>5</sup> | 606.48 (311.6) | 605.05 (323.43) | 0.97 |
| ***Post-operative hemodynamics*** |  |  |  |
| CVP, mm Hg | 8.2 (3.4) | 8.0 (3.0) | 0.50 |
| mPAP, mm Hg | 23.4 (6.5) | 22.9 (7.2) | 0.57 |
| CO, L/min | 6.0 (1.4) | 5.3 (1.0) | <0.001 |
| CI, L/min/m | 2.8 (0.5) | 2.8 (0.5) | 0.75 |
| PVR, dyn·s/cm<sup>5</sup> | 200.72 (103.62) | 230.85 (121.96) | 0.03 |

*Values are reported as mean and standard deviation unless reported otherwise. Abbreviations: RAP: right atrial pressure; mPAP: mean pulmonary artery pressure; CO: cardiac output; CI: cardiac index; PVR: pulmonary vascular resistance; CVP: central venous pressure*

Of the 500 eicosanoids analytes assayed, we found that 24 eicosanoids were increased and 6 eicosanoids were decreased in males compared to females with CTEPH after adjusting for age and BMI (**Table 3**). Notably, 8 of the 24 eicosanoids that were increased in males with CTEPH have been predominantly implicated in pro-inflammatory pathways with odds ratios ranging from 0.2 to 0.6 per 1-SD log-analyte, *p*= <10^−4^ (**Figure 1**). The potentially pro-inflammatory eicosanoids that were increased in males included leukotriene D_4_ (LTD_4_), octadecadienoic acids, and several novel eicosanoids. On the other hand, females were found to have higher levels of 6 eicosanoids, including several prostaglandins and leukotriene B_4_ (LTB_4_). Of note, none of the eicosanoids that were found to be significantly elevated in females compared to males have currently been associated with a pro-inflammatory mechanism.

**Figure 1.**
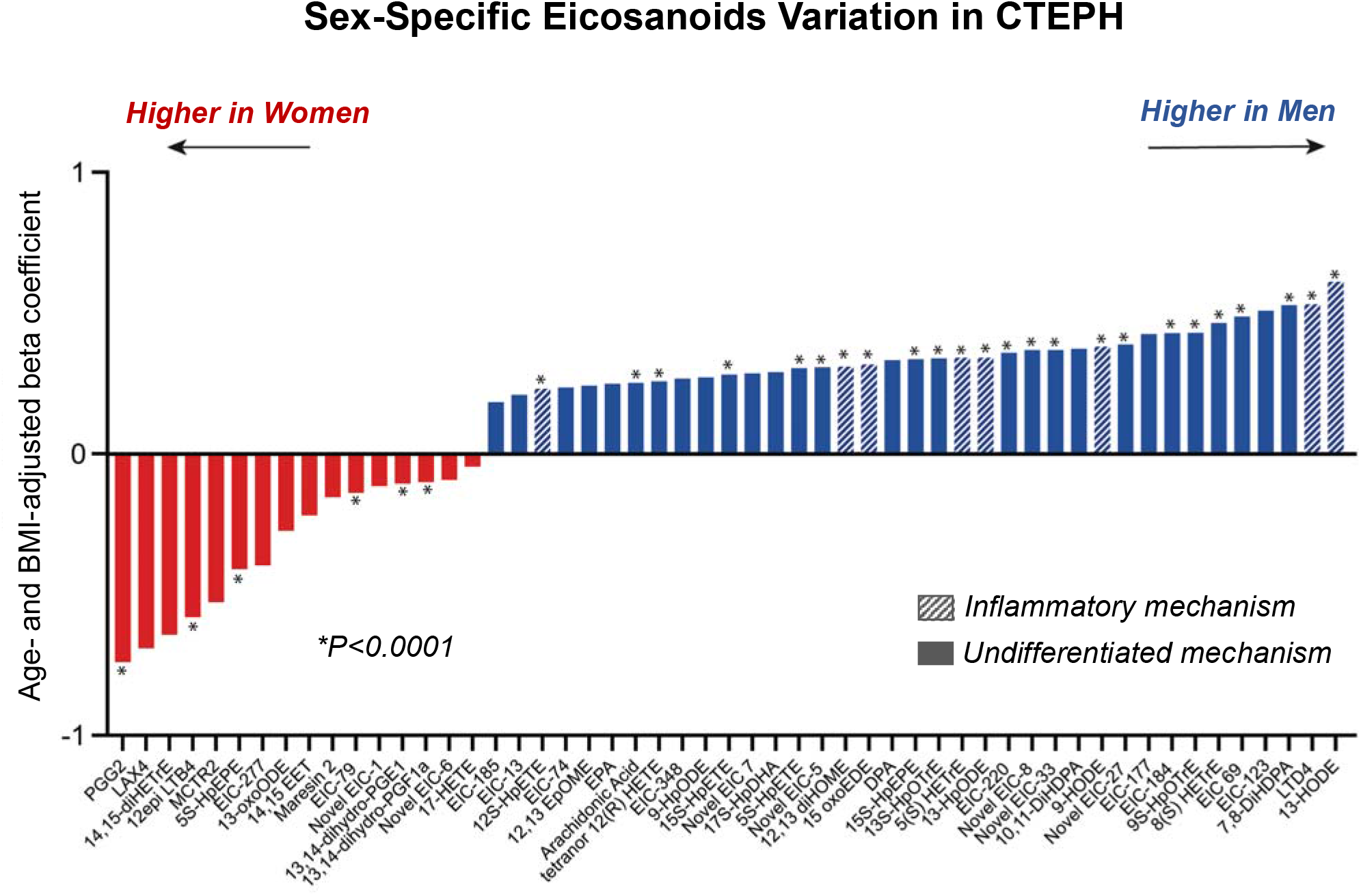
Sex-Specific Eicosanoid Variation in CTEPH. All values shown represent the difference per 1-SD increment in log-analyte level seen in men (positive values) compared women (negative values) in models adjusting for age and body mass index (BMI). Data are shown for all eicosanoids demonstrating sex-specific associations with P<10-3 and associations meeting the conservative significance threshold of P<10-4 are denoted with an asterisk. *Abbreviations: PGG2: Prostaglandin G2; 12 LTB4: 12-Leukotriene B4; 5S-HpEPE: 5-hydroperoxyicosapentaneoic acid; EIC_79: undescribed eicosanoid; 13, 14-dihydro-PGE1: 13, 14-dihydro-prostaglandin E1; 13, 14-dihydro-PGF1a: 13, 14-dihydro-prostaglandin F1a; 12S-HpETE: 12S-Hydroperoxyeicosa-tetraenoic acid; tetranor 12(R) HETE: tetranor 12 (R) -Hydroxyeicosatetraenoic acid; 15S-HpETE: 15S-Hydroperoxyeicosa-tetraenoic acid; Novel EIC_5: undescribed eicosanoid; 12, 13 diHOME: 12,13-dihydroxy-9-octadecenoic acid; 15S-HpEPE: 15S-hydroperoxy-eicosapentaenoic acid; 13S-HpOTrE: 13S-Hydroperoxyoctadecatrienoic acid; 5(S)-HETrE: 5S-Hydroxy-eicosatrienoic acid*

## DISCUSSION

In our cohort of 287 patients with CTEPH, we found significant sex-specific differences in eicosanoid profiles, including several distinct analytes previously implicated in pro-inflammatory pathways that were elevated among males. Overall, the total number of eicosanoids demonstrating significant sex-specificity after adjusting for age and BMI was substantially higher in males than females. A higher level of pro-inflammatory eicosanoids in males could at least partially contribute to variations in clinical presentations and outcomes. Importantly, there was no difference between male and female in clinical hemodynamic or UCSD surgical classification to explain the differences in eicosanoid profile. Taken together, these findings offer new insights regarding sexual dimorphism in PH and CTEPH, in particular.

The current study builds from prior predominantly experimental studies that have reported sex bias in the degree to which certain eicosanoid pathway analytes are found to be upregulated or downregulated in various settings. In general, the cyclooxygenase (COX) pathway derivatives prostaglandin E2 (PGE_2_) and thromboxane A1 (TXA_2_) have been found higher among males while prostaglandin I2 (PGI_2_) has been found higher among females in experimental models of myocardial infarction or hypertension or renovascular disease.^11^ With respect to lipoxygenase (LOX) pathway derivatives, leukotrienes tend to be produced more avidly in female compared to male neutrophils after provocation whereas levels of 5-LOX, lipoxin A1 (LXA_4_), and lipoxin B_4_ (LXB_4_) are generated to higher levels in the male than in the female left ventricle in context of experimental myocardial infarction.^11^ For cytochrome (CYP) pathway derivatives, experimental studies suggest that vascular and cardiac levels of the protective epoxyeicosatrienoic acids (EETs) are typically higher in females while hydroxyeicosatetraenoic acids (HETEs) and 20-HETE are usually higher in males.^11^

Extending from these previous experimental studies, we found in a clinical cohort of humans with manifest CTEPH that females were more likely to have higher levels of LTB_4_ in addition to a number of prostaglandin species implicated in vasoactive and thrombosis pathways. By contrast, males were significantly more likely to have higher levels of LTD_4_, octodecanoids, and a broader range of novel eicosanoids – of which a substantial number have been previously identified as central to inflammatory activity.

Limitations of our study included the single cohort and cross-sectional study design. Future investigations in separate cohorts are needed to evaluate the generalizability of our findings, and longitudinal measures would clarify the temporal associations between eicosanoids and disease phenotypes as well their treatments. Additional work is also needed to examine sex differences in eicosanoids and their relation to functional outcomes in CTEPH.^12^ Furthermore, focused analytics work is needed to clarify the exact biochemical identities of novel eicosanoids found to be associated with sex-specific CTEPH disease traits. Notwithstanding these limitations, our study had several strengths including the relatively large number of both female and male patients with CTEPH studied and the comprehensive panel of eicosanoids assayed.

In summary, we found evidence of significant and substantial sex-specific variation of circulating eicosanoids activity in patients with CTEPH, with an intriguing predominance of inflammatory pathway related eicosanoids seen in males. Given that inflammatory as well as vasoactive eicosanoid pathways can be targeted by a number of available therapeutics, our finding of sex divergent eicosanoid profiles in CTEPH suggest tractable next approaches for not only improving our understanding of CTEPH pathophysiology but also for developing even more tailored treatments based on potential sex-specific disease mechanisms.

## Data Availability

Requests for de-identified data may be directed to the corresponding authors (MJ, SC) and will be reviewed by the Office of Research Administration at UCSD prior to issuance of data sharing agreements. Data limitations are designed to ensure patient and participant confidentiality.

## Non-standard Abbreviations and Acronyms

BMI: Body mass index
CTEPH: Chronic thromboembolic pulmonary hypertension
PAH: Pulmonary arterial hypertension
LC-MS: Liquid chromatography - mass spectrometry
mRAP: Mean right atrial pressure
PAH: Pulmonary arterial hypertension
PVR: Pulmonary vascular resistance
RHC: Right heart catheterization
6MWD: 6-minute walk distance

## ACKNOWLEDGMENTS

We thank contributors who collected samples used in this study, as well as patients and their families, whose help and participation made this work possible.

## AUTHOR CONTRIBUTION

MA, SC, MJ and NHK conceived and designed the overall study. MA, VP, MM and TF acquired the data. MA, JDW, ABT, GAM and SC conceived the analysis of the study and analyzed the data. MA, ABT, GAM and SC drafted the manuscript, and all authors edited the manuscript.

